# Semantic Drift and Reporting Stability in VAERS: An Embedding-Based Analysis of Vaccine Safety Narratives

**DOI:** 10.1101/2025.11.10.25339934

**Authors:** David Rosado

## Abstract

The Vaccine Adverse Event Reporting System (VAERS) is critical for post-market vaccine surveillance to understand the progression of rare but severe adverse events. Traditional analysis uses event frequencies and odds ratios to determine safety signals focusing on biological causality. Linguistic reports provide underused behavioral information that affect reporting dynamics. This study aims to quantify temporal and semantic drift in vaccine safety narratives and examine its relationship to epidemiological reporting patterns. The narrative text for Influenza, MMR and Pneumococcal vaccines were preprocessed and converted to numerical format using embedding models to quantify similarity between reports. The cosine similarity distance was used to calculate a distortion index, a composite measure of linguistic heterogeneity and volatility. The index was correlated with behavioral indicators as text length, reporting lag and distributional changes in reported symptoms and tested for robustness using bootstrap sampling. Additionally, semantic regimes were found using changepoint detection, showing that language drift can act as a measurable signal for the behavioral dynamics of pharmacovigilance.

## Introduction

The Centers for Disease Control and Prevention (CDC) and the U.S. Food and Drug Administration (FDA) jointly operate the Vaccine Adverse Event Reporting System (VAERS), a national passive surveillance program established in 1990 to monitor adverse events following immunization [3]. VAERS accepts reports from healthcare providers, manufacturers, and the public, serving as an early-warning system for the detection of potential vaccine-safety signals in phase 4 trials, even with well-recognized limitations introduced by voluntary submissions [1, 2].

Traditional vaccine-safety surveillance has focused primarily on quantitative disproportionality metrics (PRR, ROR) which quantify what, but not how it is reported [10]. Despite extensive quantitative monitoring of vaccine safety data, current pharmacovigilance frameworks are largely blind to changes in reporting behavior, shifts in how individuals describe, prioritize, or frame adverse events. As a result, variations in language use, which reflect public perception, clinical framing, and temporal attention, remain unmeasured sources of uncertainty in vaccine-safety surveillance.

This gap has important implications on the interpretation of VAERS data because reporting behavior itself evolves in response to external stimuli: media coverage, regulatory announcements, or shifts in population demographics. Hence, if the linguistic structure of reports changes, so does the interpretability of signal metrics built on them. Without tools to quantify these semantic dynamics, systems like VAERS cannot distinguish between true safety signals and behavioral or cognitive drift in reporting.

VAERS free-text narratives contain rich information on the reporters’ own descriptions of symptoms, contexts, and experiences, encoding behavioral information about how the system is used, how awareness and concern evolve, and how events are framed across time [1]. Shifts in descriptive language can thus reveal evolving reporting practices or emerging narratives - phenomena that cannot be measured using count-based analyses [9, 4].

Although natural language processing (NLP) techniques have recently been applied to vaccine adverse-event data, most efforts have emphasized classification or feature extraction rather than temporal or semantic analysis of the free-text [1, 5]. In other domains of NLP and social media analytics, the concept of semantic drift, the gradual change of word or concept meanings over time, has been used to detect evolving discourse, attention, and framing [9, 4]. In the context of vaccine safety surveillance, this invites the hypothesis that the semantics of how adverse events are described in VAERS may shift with external circumstances: new vaccine formulations, heightened media coverage, or changes in the demographics of reporters. The linguistic shifts in how adverse events are conceptualized and expressed may, in turn, signal changes in reporting behavior or broader system dynamics [1, 5].

This study introduces an embedding-based metric, the Semantic Distortion Index (*ζ*), designed to quantify two dimensions of narrative change: (i) language heterogeneity (dispersion, *δ*) and (ii) semantic volatility (instability, *ι*). The index is computed for three vaccine groups (Influenza, MMR, and Pneumococcal) to facilitate comparison of narrative dynamics across distinct reporting populations. The association between this index and key epidemiological covariates - reporting lag, narrative length, symptom counts, Shannon entropy, and Jensen-Shannon divergence (JSD) - is then evaluated, with estimator robustness assessed through bootstrap resampling. Change-point analysis is additionally applied to identify persistent shifts in semantic behavior over time.

## Methods

This analysis was restricted to reports received between 2010 and 2019 to ensure a decade of consistent electronic formatting, sufficient narrative length, and representation of vaccines. Earlier records were excluded due to sparse text and inconsistent MedDRA coding, and reports from 2020 onward were excluded due to the extraordinary surge in COVID-19 related submissions, changes in reporting behavior, and media-driven linguistic shifts that merit a separate study.

Influenza, MMR, and Pneumococcal vaccines were selected because they provide long, continuous reporting histories with sufficient monthly volume to support reliable embedding-based linguistic estimates; vaccines with sparse or intermittent reporting could produce unstable estimators. These three vaccine groups also represent distinct reporting populations: Influenza is a high-volume seasonal vaccine primarily administered to adults, MMR represents routine pediatric immunizations, and Pneumococcal vaccines are predominantly administered to older adults. This diversity enables meaningful behavioral comparisons across reporting contexts. In addition, embedding 2-3 million narratives across all available vaccines would be computationally prohibitive. Restricting the analysis to three representative vaccine classes allows for detailed assessments while keeping computation tractable.

For each report, the free-text narrative was processed with the Python library Polars [8] to remove null values, whitespace, and redundant text, and expand common clinical abbreviations to full text, such as “htn” to hypertension. Subsequently the embedding for each symptom narrative was computed using OpenAI’s large embedding model (text-embedding-3-large) to preserve semantic richness and nuance, transforming each text to a numerical vector of 3072 dimensions [7]. The cosine similarity was computed for each pair of embeddings to establish semantic proximity between reports, because in high dimensional space semantically close text points in similar directions regardless of embedding norms [6].

Principal Component Analysis (PCA) was applied to project the original high dimensional embedding space to the two directions of maximum variance to aid with visualization of the linguistic variability. Visualizations were created by partitioning the space in a 60 x 60 grid and measuring the relative density on each bin. While this method does not preserve the high dimensional structure of the data, it enables a qualitative assessment of changes in density and distribution of the linguistic space. When inspecting yearly embeddings of VAERS text in the low-dimensional approximation space, two empirical patterns emerged, 1) reports varied widely in how they described similar events, and 2) internally consistent clusters shifted their position in embedding space.

These observations suggested that VAERS could be conceptualized as a dynamic linguistic system with vocabulary and framing conventions that evolve over time. Under the assumption that reports described comparable clinical events, it was hypothesized that such behavioral and perceptual changes did not necessarily alter frequencies, rather manifested structural shifts in the linguistic space. If descriptions became more linguistically diverse or heterogeneous, the population’s shared representation of “the vaccine adverse event” fragmented, representing a form of semantic entropy. On the other hand, even when language remained internally consistent, its the center of gravity could drift over time, reflecting temporal volatility in collective framing.

To operationalize these constructs, two measurable components were constructed using the cosine similarities: dispersion (*δ*) and instability (*ι*). Dispersion was defined as the interquartile range of pairwise cosine similarities between all narrative embeddings within each month. Using the interquartile range rather than standard deviation ensured robustness to outlier embeddings, which may arise from data quality issues in patient-reported narratives. Semantic instability was described as the absolute month-to-month change in median cosine similarity to capture shifts in typical report characteristics, while remaining robust to extreme values. Because both are on different numeric scales but conceptually additive contributors to “semantic distortion,” each was standardized (z-scored) and combined additively to define the Semantic Distortion Index (*ζ*).

Formally, let 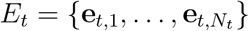 denote the set of narrative embeddings for all reports *N*_*t*_ in month *t*, hence the cosine similarity between reports *I* and *j* in month *t* is expressed as:

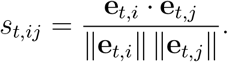

### Semantic Dispersion

Semantic dispersion is defined as the interquartile range (IQR) of cosine similarities *s*_*t,ij*_:

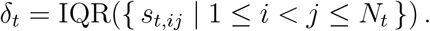

### Semantic Instability

Let *M*_*t*_ = median({*s*_*t,ij*_ | 1 ≤ *i* < *j* ≤ *N*_*t*_ }) be the monthly median similarity. Temporal instability is then

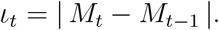

### Semantic Distortion

After z-score normalization of *δ*_*t*_ and *ι*_*t*_ across months,

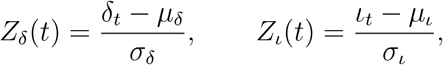

the semantic distortion index is defined as

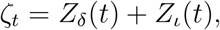

which represents the overall deviation of the system from its baseline linguistic equilibrium, a composite measurement for semantic stability.

Bootstrapping was employed for two complementary purposes: (1) random subsampling to mitigate computational costs of pairwise similarity calculations in high-dimensional embedding space, and (2) estimation of 95 % confidence intervals (CIs) and learning curves for correlation coefficients. Ten random seeds and three monthly sample sizes (N=100,200,300) were used, resulting in 30 independent bootstrap runs. Confidence intervals were computed using non-parametric percentile bands

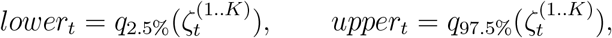

and seed agreement were derived by calculating the Spearman’s Rank correlation coefficient (*ρ*) for each sample size across the 10 seeds for each *N* .

Furthermore, the relationship between linguistic variability measured by *ζ*, and reporting behavior was assessed using the Spearman’s rank correlation coefficient. Correlations were computed between the monthly distortion index and five epidemiological or narrative covariates: average reporting lag (days between vaccination and report submission), average narrative text length, number of unique symptom terms, the Shannon entropy, and Jensen–Shannon divergence (JSD) applied on symptom distributions with two times the standard error (2SE). Correlations were computed on data detrended with the Seasonal-Trend decomposition using LOESS algorithm (STL), using the seasonal and residual components to measure variation dominated by short to medium drift. Statistical significance over all fifteen hypothesis (5 covariates and 3 vaccines) was evaluated via two-sided tests with Benjamini–Hochberg correction (*q* < 0.05).

The mean monthly distortion across bootstrap replicates was analyzed to identify structural regime shifts in linguistic behavior. Series were deseasonalized with STL retaining trend and residual components, and subjected to changepoint detection using the Pruned Exact Linear Time (PELT) algorithm with an *L*_2_ cost function and Bayesian Information Criterion (BIC) penalty (log *n*). Autocorrelation analysis confirmed residual independence, validating changepoint assumptions.

## Results

At the time of analysis, the VAERS database contained 3,599,087 total reports, of which approximately 81.7% included narrative text suitable for natural language processing. Among these, Influenza, MMR, and Pneumococcal vaccines accounted for 299,679, 128,841, and 189,241 reports, respectively. After applying inclusion criteria, restricting to records with non-empty narratives and vaccination dates between 2010 and 2019, the final analytic datasets comprised 168,768 Influenza, 42,392 MMR, and 89,567 Pneumococcal reports.

Projection of the high-dimensional embedding space onto the first two principal components by applying PCA revealed temporal shifts in density, suggesting gradual redistribution of reports along the dominant variance directions. Although the two-dimensional representation cannot generalize to the complete high-dimensional structure, it effectively illustrates localized changes in semantic composition across years (Figure 1) .

**Figure 1.**
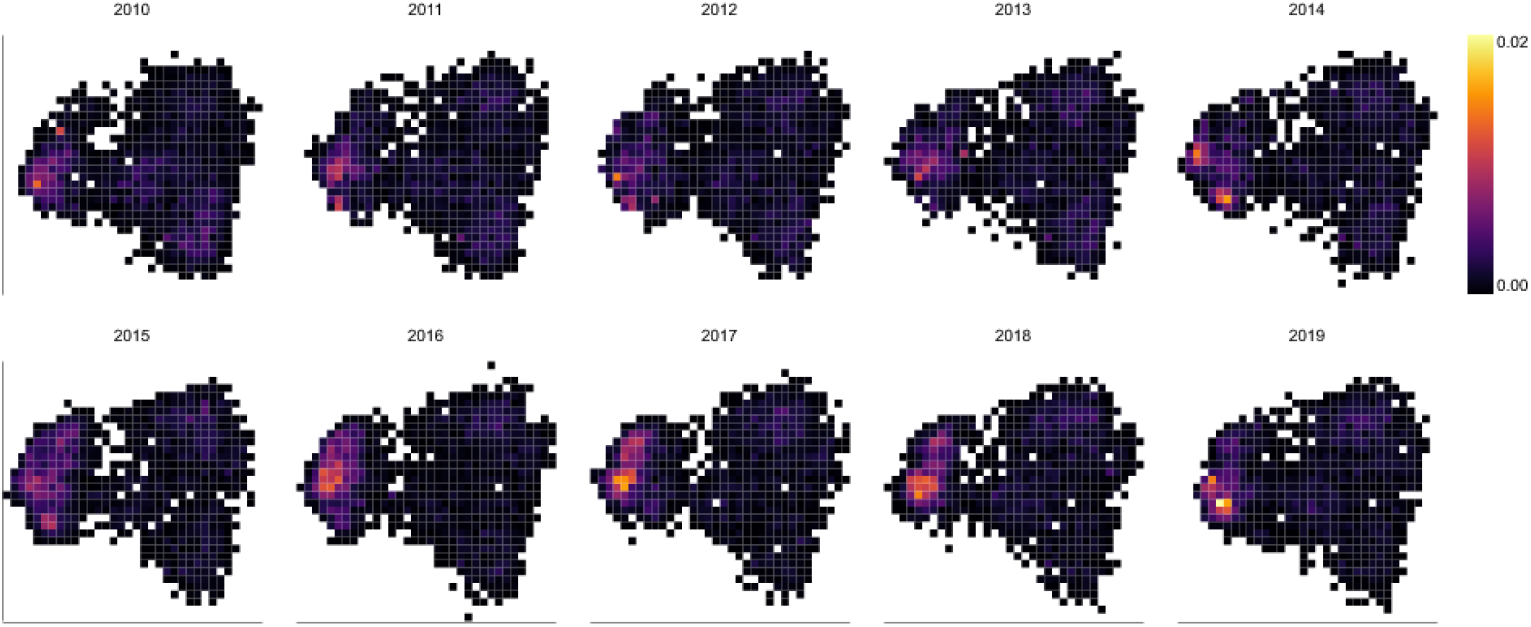
Principal Component Analysis. Heatmap illustrating the two-dimensional projection of the embedding space for Influenza vaccines, 2010 - 2019. Density represents frequency count per bin in a 60 x 60 grid.

The temporal profiles of the distortion index revealed distinct dynamic patterns across vaccine groups. Influenza narratives exhibited marked seasonality, with recurring annual peaks aligning with the vaccination cycle. MMR reports showed a clear regime shift around 2014, suggesting a structural change in linguistic behavior, whereas Pneumococcal vaccines displayed overall stability with mild short-term volatility (Figures 2–4).

**Figure 2.**
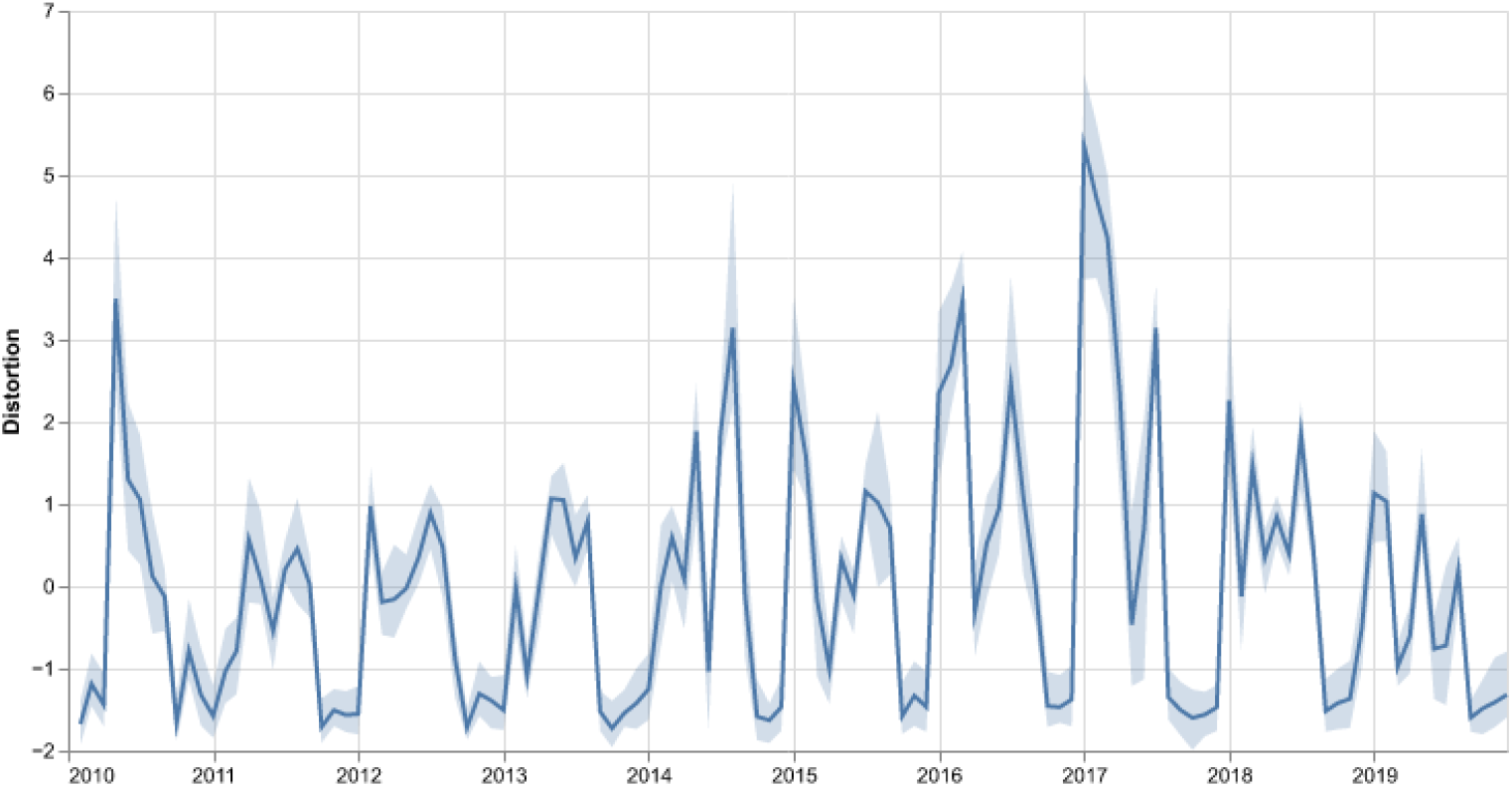
Semantic Distortion Index. Line plot and 95% confidence intervals for the Influenza vaccines’ Semantic Distortion Index computed monthly from 2010 to 2019.

**Figure 3.**
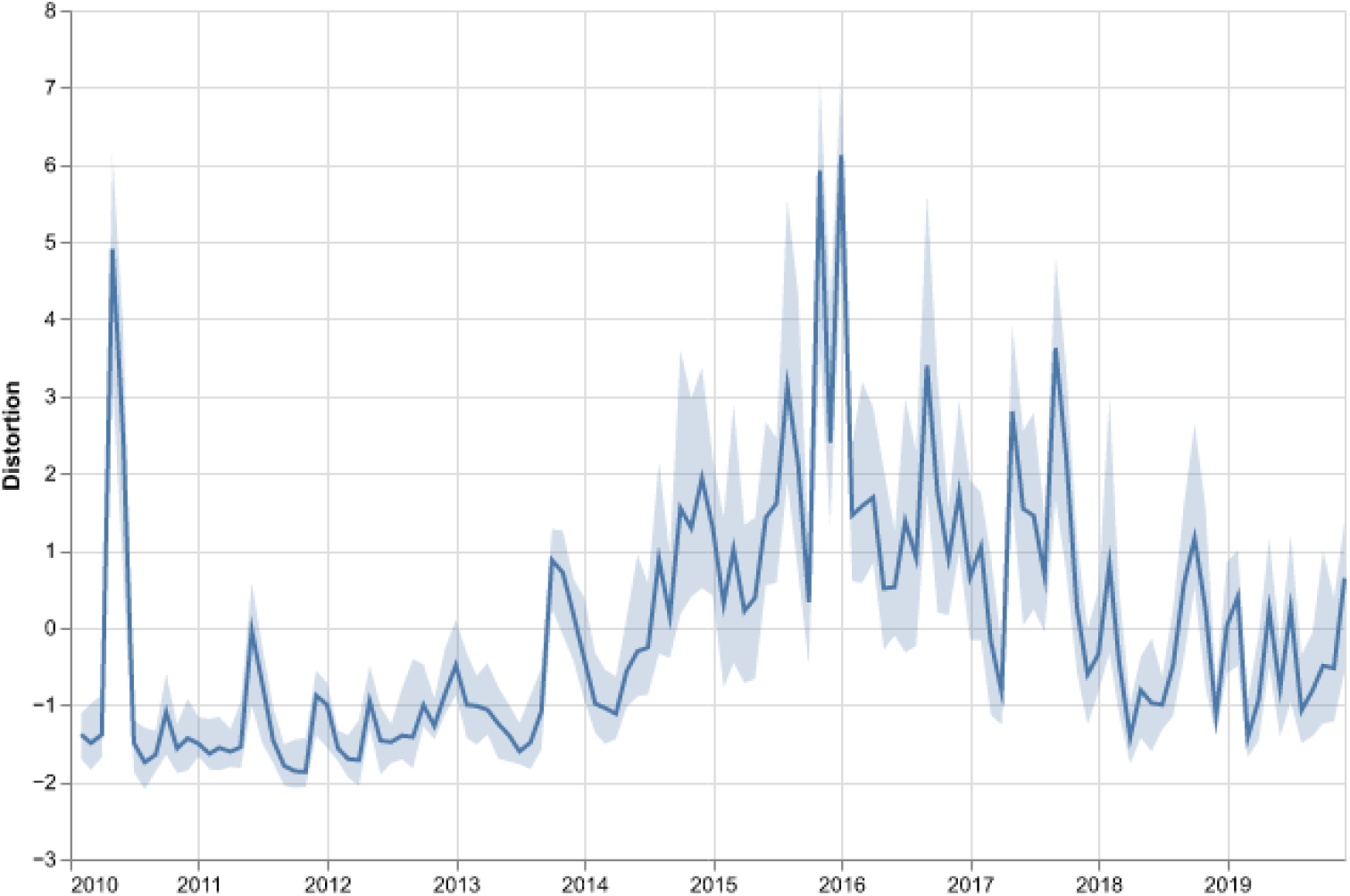
Semantic Distortion Index. Line plot and 95% confidence intervals for the MMR vaccines’ Semantic Distortion Index computed monthly from 2010 to 2019.

**Figure 4.**
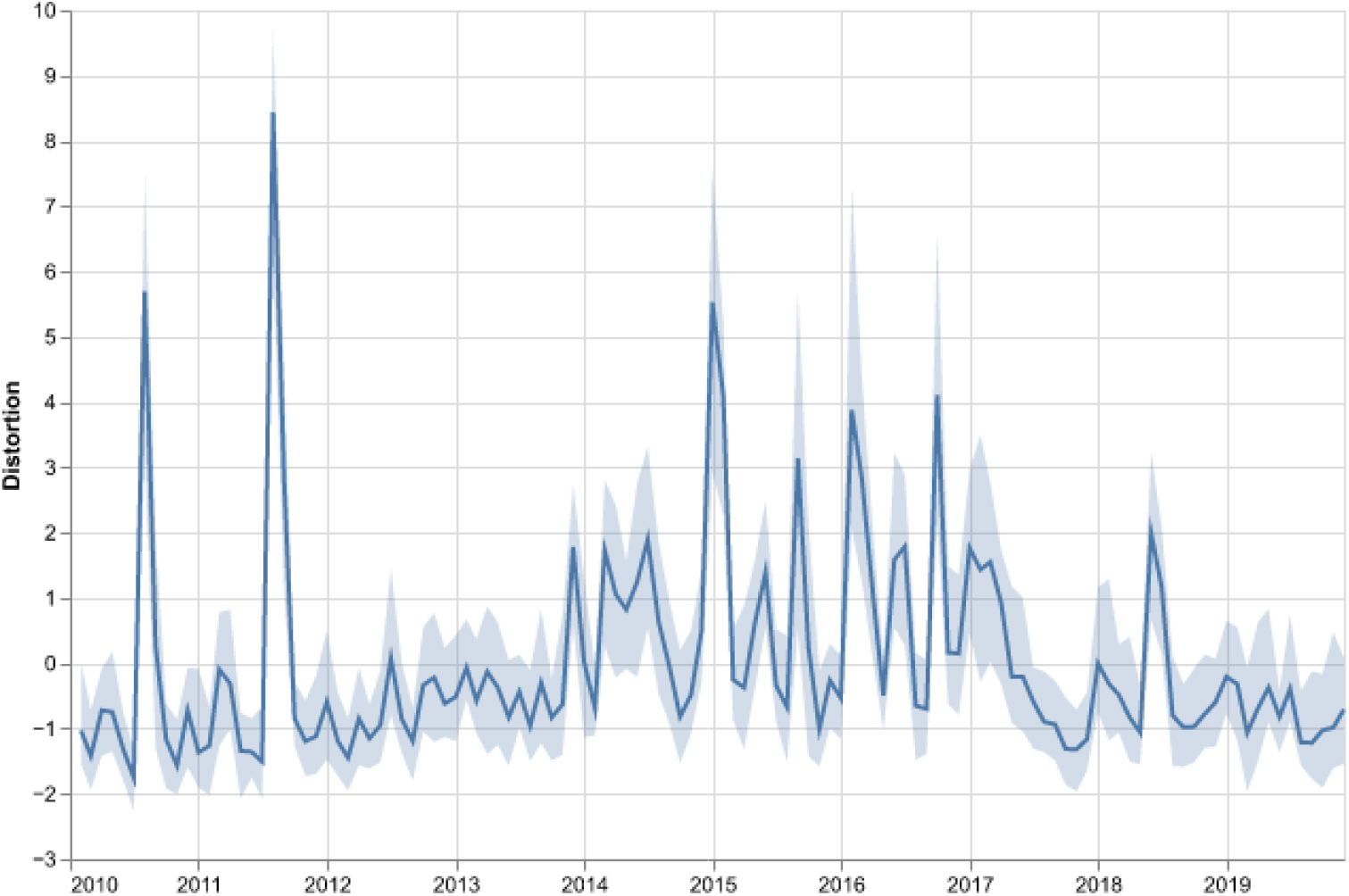
Semantic Distortion Index. Line plot and 95% confidence intervals for the Pneumococcal vaccines’ Semantic Distortion Index computed monthly from 2010 to 2019.

The distortion index in Influenza vaccines exhibited strong positive correlations with reporting lag (*ρ* = 0.68, *p* < 10^−16^) and narrative length (*ρ* = 0.60, *p* < 10^−12^), suggesting that delayed and longer reports tend to show greater linguistic variability. Moderate correlations with JSD and report volume indicate partial alignment with distributional drift and reporting intensity (Table 2).

**Table 1:** Definitions of semantic descriptors. Mathematical definitions and their corresponding interpretations as language-based metrics.

| Concept | Metric | Captures |
| --- | --- | --- |
| Dispersion | IQR of pairwise cosine similarities between all narrative embeddings | Semantic heterogeneity, diversity of wording and conceptual framing within that period |
| Instability | Absolute month-to-month change in median cosine similarity | Temporal volatility, how rapidly the center of linguistic meaning moves |
| Distortion | Composite of dispersion and instability | Semantic stress, how far the system deviates from linguistic equilibrium |

**Table 2:** Spearman Rank Correlation. Correlation coefficients between the Semantic Distortion Index for Influenza vaccines and epidemiological covariates. Estimates shown with 2SE; p-values are FDR-adjusted.

| Variable | $\rho \pm 2SE$ | Adjusted p | Significant |
| --- | --- | --- | --- |
| Reporting lag | $0.68 \pm 0.011$ | 6.83e-17 | Yes |
| Narrative length | $0.60 \pm 0.009$ | 2.31e-13 | Yes |
| JSD | $0.52 \pm 0.019$ | 3.09e-10 | Yes |
| Shannon entropy | $0.21 \pm 0.020$ | 1.34e-2 | Yes |
| AE counts | $0.25 \pm 0.017$ | 6.27e-3 | Yes |

Conversely the distortion index in MMR vaccines displayed a weak relationship with reporting lag and narrative length. Correlations were not found on JSD, adverse event unique count and Shannon Entropy, indicating that distortion captures qualitative changes in linguistic organization, rather than broad lexical shifts or increases in vocabulary diversity (Table 3).

**Table 3:** Spearman Rank Correlation. Correlation coefficients between the Semantic Distortion Index for MMR vaccines and epidemiological covariates. Estimates shown with 2SE; p-values are FDR-adjusted.

| Variable | $\rho \pm 2SE$ | Adjusted p | Significant |
| --- | --- | --- | --- |
| Reporting lag | $0.20 \pm 0.016$ | 4.92e-2 | Yes |
| Narrative length | $0.30 \pm 0.016$ | 1.13e-3 | Yes |
| JSD | $0.20 \pm 0.019$ | 6.62e-1 | No |
| Shannon entropy | $-0.31 \pm 0.023$ | 1.13e-3 | Yes |
| AE counts | $-0.17 \pm 0.025$ | 9.76e-2 | No |

Pneumococcal vaccines showed a moderate positive correlation to reporting lag (*ρ* = 0.43, *p* < 10^− 5^ and narrative length (*ρ* = 0.31, *p* < 10^− 4^, and no association with distributional changes in adverse events, also indicating qualitative linguistic organization, rather than changes in vocabulary diversity (Table 4).

**Table 4:**
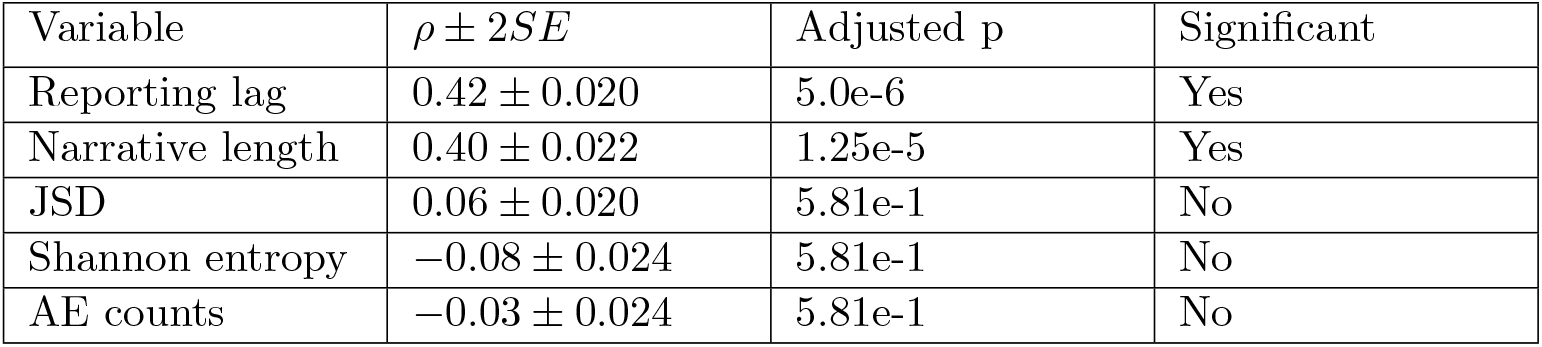
Spearman Rank Correlation. Correlation coefficients between the Semantic Distortion Index for Pneumococcal vaccines and epidemiological covariates. Estimates shown with 2SE; p-values are FDR-adjusted.

| Variable | $\rho \pm 2SE$ | Adjusted p | Significant |
| --- | --- | --- | --- |
| Reporting lag | $0.42 \pm 0.020$ | 5.0e-6 | Yes |
| Narrative length | $0.40 \pm 0.022$ | 1.25e-5 | Yes |
| JSD | $0.06 \pm 0.020$ | 5.81e-1 | No |
| Shannon entropy | $-0.08 \pm 0.024$ | 5.81e-1 | No |
| AE counts | $-0.03 \pm 0.024$ | 5.81e-1 | No |

Across all vaccine groups, reporting lag and narrative length showed consistent positive correlations with distortion after FDR correction, indicating that longer and more delayed reports tend to exhibit greater linguistic drift. Shannon entropy was significant in Influenza and MMR but not in Pneumococcal reports, suggesting that lexical diversity contributes to distortion primarily in linguistically richer datasets. Jensen–Shannon divergence and adverse-event count were significant only for Influenza, indicating that this group exhibits both large-scale lexical reorganization and changes in reported AE structure alongside distortion. These patterns were stable across bootstrap replicates and sample sizes, and after adjusting p-values with Benjamini–Hochberg procedure (*α* = 0.05) on the median p-value.

Changepoint analysis of the detrended distortion trajectories identified multiple linguistic regimes across vaccine types. Influenza exhibited three phases marked by transitions in mid-2014, early-2017, and mid-2017, corresponding to shifts in the seasonal distortion baseline. MMR displayed a more fragmented structure with seven regime shifts between 2014 and 2018, suggesting recurrent reorganizations in its linguistic dynamics. Pneumococcal reports remained comparatively stable, with only two changepoints, around early-2014 and mid-2017, indicating fewer disruptions in reporting language (Figures 5 - 7).

**Figure 5.**
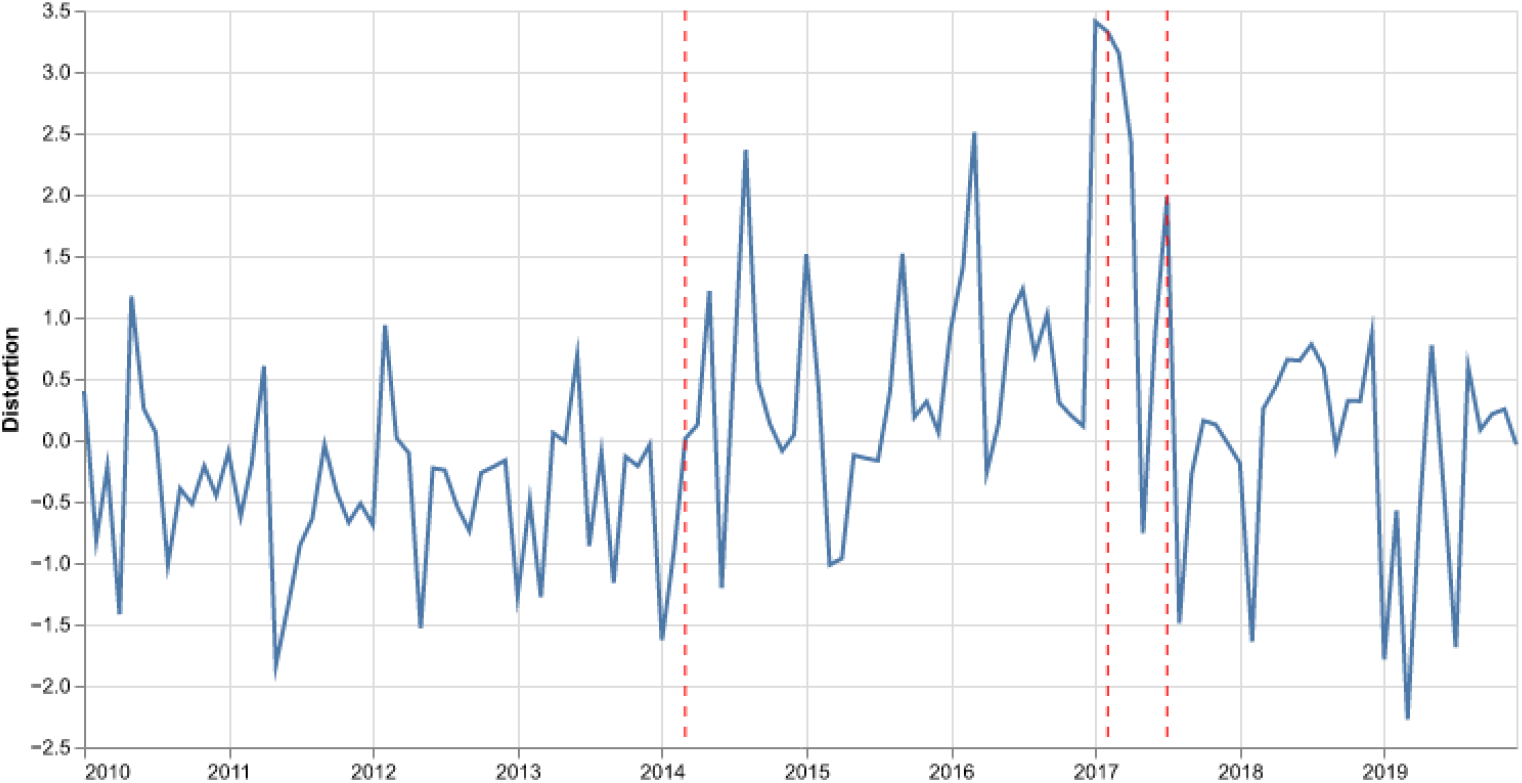
Changepoint detection analysis. Semantic Distortion Index after STL detrending for Influenza vaccines, 2010–2019. Changepoints detected in early 2014, early 2017, and mid-2017.

**Figure 6.**
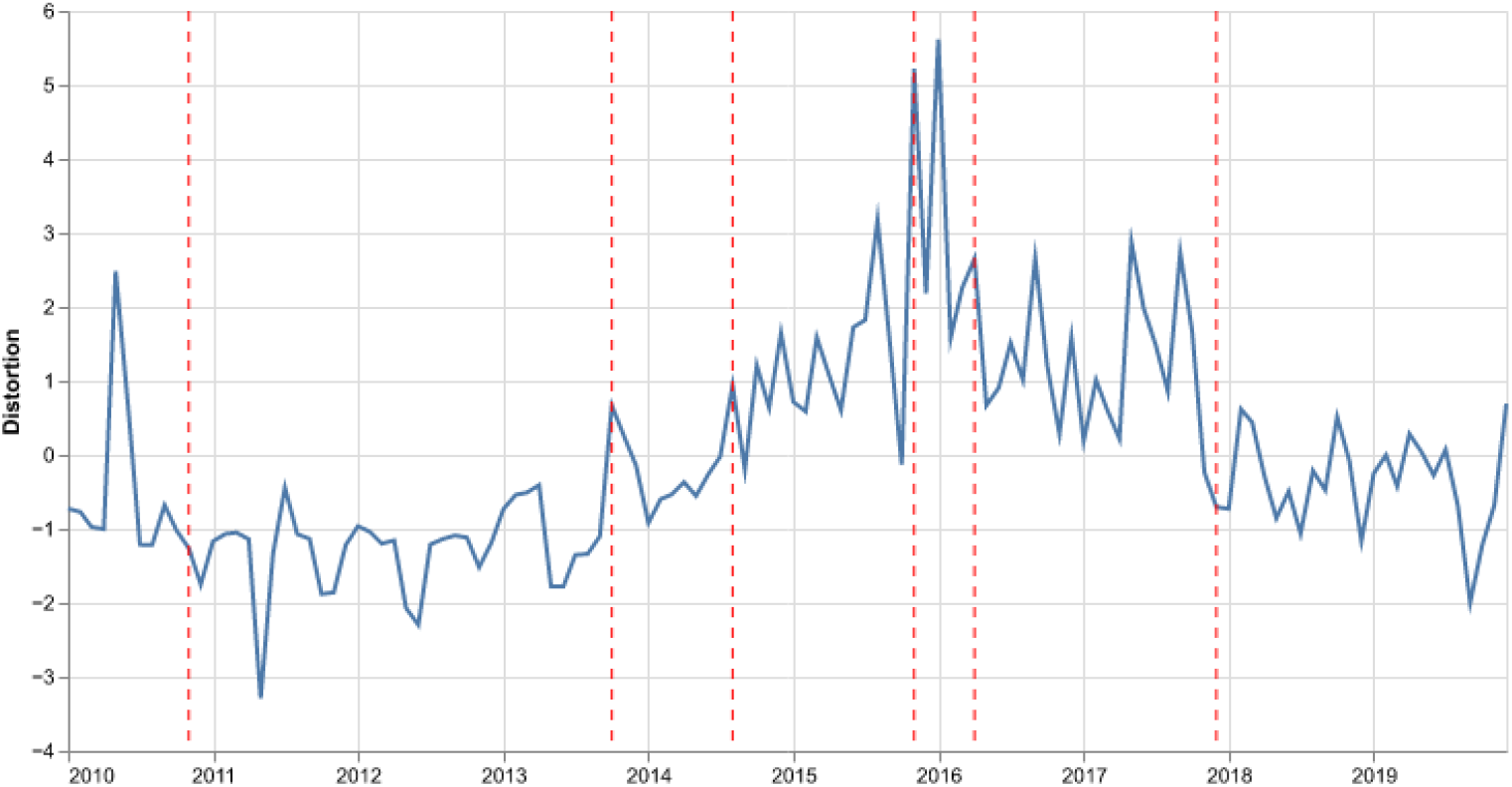
Changepoint detection analysis. Semantic Distortion Index after STL detrending for MMR vaccines, 2010–2019. Changepoints detected in late 2010, late 2013, mid-2014, late 2015, early 2016, and late 2017.

**Figure 7.**
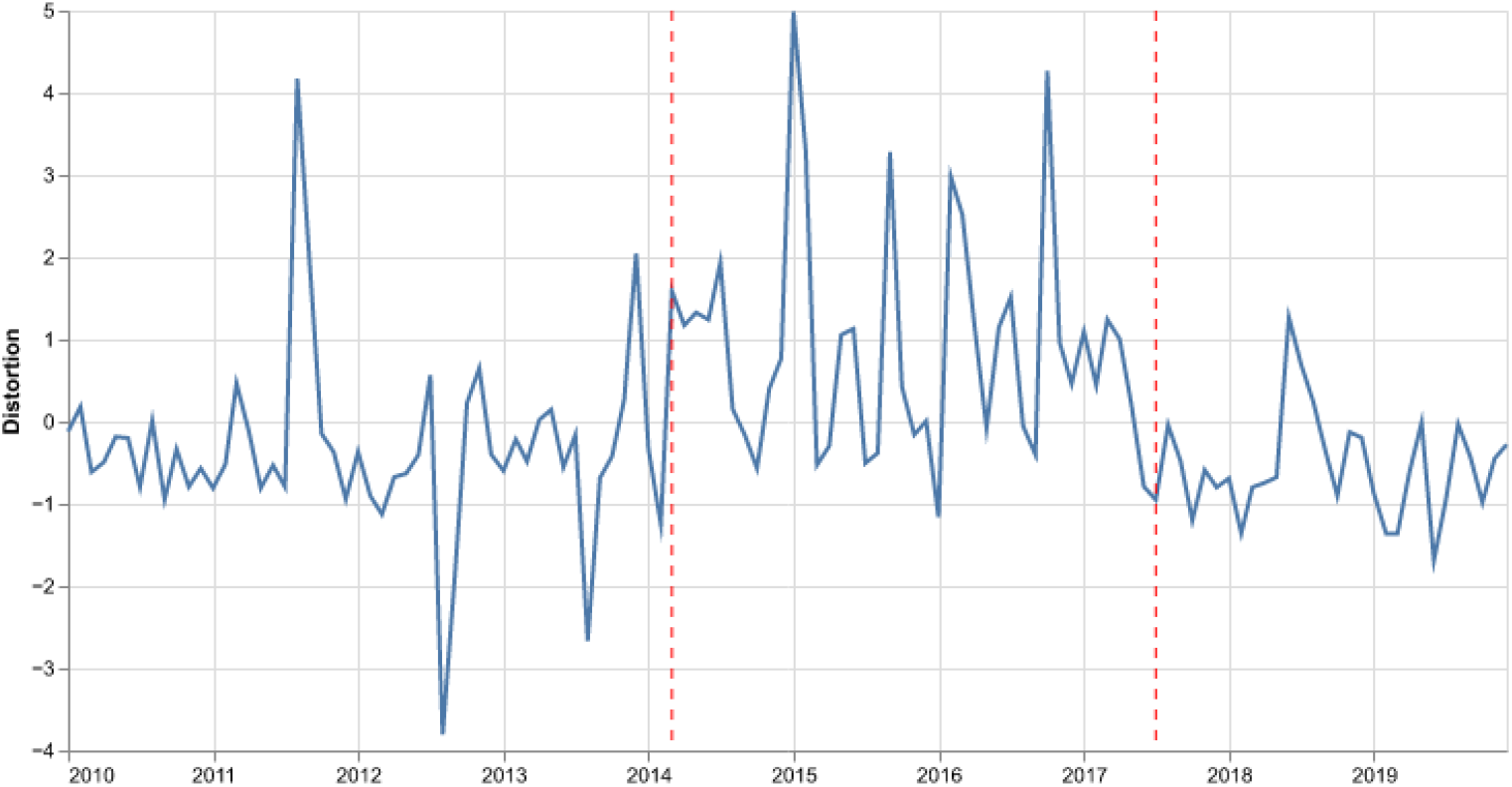
Changepoint detection analysis. Semantic Distortion Index after STL detrending for Pneumococcal vaccines, 2010–2019. Changepoints detected in early 2014 and mid-2017.

Seed agreement in bootstrap estimates increased monotonically with sample size across vaccines, indicating high reproducibility of the distortion index (Table 5). Volume and temporal continuity in Influenza and MMR lead to stable statistics for the distortion index, contrary to Pneumococal vaccines with sparser reporting and smaller monthly volumes.

**Table 5:** Seed agreement. Bootstrap estimates across ten seeds for sample sizes of 100, 200, and 300. Estimates shown with 2SE.

| Vaccine | N=100 | N=200 | N=300 |
| --- | --- | --- | --- |
| Influenza | $0.89 \pm 0.017$ | $0.95 \pm 0.008$ | $0.97 \pm 0.006$ |
| MMR | $0.83 \pm 0.024$ | $0.95 \pm 0.007$ | $0.98 \pm 0.004$ |
| Pneumococcal | $0.62 \pm 0.056$ | $0.79 \pm 0.031$ | $0.85 \pm 0.023$ |

## Discussion

The three semantic descriptors, dispersion, instability, and distortion provide a multidimensional view of how linguistic behavior in VAERS narratives reflects collective cognition and information flow in vaccine safety reporting. Dispersion captures the degree of linguistic diversity across reports. Elevated dispersion suggests fragmentation in how individuals articulate adverse events, potentially driven by heightened uncertainty, media amplification, or the introduction of new vaccines or syndromic patterns. In such periods, multiple linguistic frames may coexist as reporters draw from heterogeneous sources of information to describe their experiences. Conversely, low dispersion indicates linguistic convergence, where the reporting language becomes more standardized and cognitively routinized, reflecting shared understanding or institutional reinforcement of specific diagnostic or narrative templates.

Instability represents temporal volatility in the underlying semantic structure. High instability denotes moments when the semantic embedding space reorganizes rapidly; for instance when language conventions shift, clusters merge or dissolve, and conceptual relationships are renegotiated. Such episodes alter how reporters conceptualize causality or symptom patterns. Low instability, by contrast, marks linguistic inertia, where the framing of events remains consistent over extended periods, suggesting stable cognitive schemas and reporting norms.

Distortion integrates both dimensions as a proxy for semantic stress within the system. It signals moments when the collective language of reporting reconfigures under informational pressure. High distortion can therefore be interpreted as a behavioral indicator of communicative strain or epistemic uncertainty, while low distortion reflects equilibrium in the narrative ecosystem; a state in which linguistic practices, interpretive frames, and reporting incentives align coherently.

Furthermore, the strong positive association after FDR correction between distortion and reporting lag and narrative length in influenza suggests that linguistic instability increases when reports are filed long after vaccination, potentially reflecting retrospective or secondhand accounts. The weaker correlations in MMR and pneumococcal vaccines indicate more standardized reporting, consistent with pediatric and geriatric reporting forms completed by healthcare providers rather than patients. Collectively, these results show that the semantic organization of VAERS narratives is not stationary and-sensitive to population dynamics.

For influenza particularly the distortion index was correlated not only with behavioral variables (reporting lag, narrative length) but also with distributional changes in adverse events, implying that linguistic reorganization coincides with structural changes in AE co-reporting. Such shifts may bias disproportionality-based signal detection metrics that assume lexical and reporting stability. Incorporating distortion as a covariate or normalization factor could therefore improve the robustness of pharmacovigilance models, especially in periods of rapid linguistic drift or changing public discourse.

Between 2014 and 2017, the distortion trajectories of all vaccine groups showed evidence of a common period of linguistic turbulence. Although the exact timing of changepoints varied - early 2014 and mid-2017 for influenza, mid-2014 and mid-2017 for pneumococcal, and multiple transitions from late 2013 through late 2017 for MMR. The clustering of regime shifts within this interval suggests a broader systemic reorganization in the VAERS linguistic landscape rather than vaccine-specific anomalies. This period coincided with major sociotechnical transformations in pharmacovigilance: the increasing adoption of online submission portals, growing public debate over vaccine safety, and intensified digital circulation of vaccine-related narratives. Such changes likely altered not only reporting volume but also the linguistic conventions through which adverse events were described.

Within this broader transformation, each vaccine class exhibited distinct linguistic responses. Influenza and pneumococcal vaccines (primarily adult, seasonal, or preventive immunizations) showed relatively few changepoints and faster recovery toward a stable baseline. For influenza, the 2014 shift may correspond to the low-effectiveness H3N2 season, which drew public and media attention, while the 2017 transition coincided with both the rollout of VAERS 2.0 and another severe influenza season. Pneumococcal changepoints near 2014 align with the expansion of ACIP recommendations to include PCV13 for older adults, representing an institutional change in both exposure and reporting patterns. Despite these perturbations, both vaccines retained overall linguistic cohesion, consistent with reports written largely by healthcare professionals using established clinical vocabulary.

In contrast, MMR exhibited a more fragmented and volatile distortion profile, with five distinct linguistic regimes emerging between late 2013 and late 2017. This volatility overlaps with renewed public debate over childhood immunization, triggered by high-profile measles outbreaks (2014-2015) and the concurrent amplification of vaccine skepticism in online spaces. The resulting informational environment, characterized by politicization and uncertainty likely diversified the language used to describe similar adverse events. MMR narratives thus appear to capture the imprint of shifting social discourse more strongly than the adult vaccine groups, consistent with a population where parents and non-professional reporters contribute more directly to narrative content.

It is important to emphasize that these interpretations are qualitative and exploratory, not causal claims. The observed linguistic regimes likely reflect an interplay of structural, social, and behavioral influences ranging from system-level transitions in reporting infrastructure to shifts in media attention and public discourse, rather than any single determinant. The goal here is to outline plausible mechanistic pathways through which reporting practices evolve, acknowledging that confirmation of causality would require an entirely different study design, integrating controlled covariates and formal causal inference.

From a pharmacovigilance perspective, the analysis illustrates how metrics for language variation can complement traditional signal-detection methods. Monitoring linguistic stability over time may help identify periods when the reporting ecosystem itself is undergoing stress - whether due to policy change, public controversy, or data-system updates - allowing analysts to contextualize signals within broader communicative dynamics. In this sense, semantic monitoring serves as an early indicator of epistemic instability in reporting. Recognizing these qualitative shifts enriches interpretation of spontaneous-report data, helping distinguish true pharmacological signals from socially driven perturbations in language and attention.

## Conclusion

These findings suggest that linguistic dynamics in VAERS, captured through the semantic distortion index, encode behavioral and cognitive aspects of vaccine safety reporting. Periods of high distortion coincided with changes in narrative length, reporting lag, and diversity of symptoms, implying that shifts in how adverse events are described accompany shifts in how and when they are reported. This highlights the potential for embedding-based linguistic monitoring to complement quantitative signal detection methods, offering insight into changes in public attention, medical framing, or reporting consistency. Beyond VAERS, the approach establishes a framework for integrating natural language indicators into pharmacovigilance pipelines, transforming narrative text from a passive artifact into an active source of behavioral signal that is observable and quantifiable.

## Data Availability

All data produced are available online at https://vaers.hhs.gov/data/datasets.html

## References

[1] Taxiarchis Botsis et al. “Vaccine adverse event text-mining system for extracting key clinical features from VAERS narratives”. In: Journal of the American Medical Informatics Association 19.5 (2012), pp. 631– 638. doi: 10.1136/amiajnl-2011-000750.

[2] Centers for Disease Control and Prevention. Surveillance Manual, Chapter 21: Vaccine Adverse Event Reporting System (VAERS). Available at https://www.cdc.gov/surv-manual/php/table-of-contents/chapter-21-vaers.html. 2023.

[3] Centers for Disease Control and Prevention. Vaccine Adverse Event Reporting System (VAERS) Overview. Available at https://www.cdc.gov/vaccine-safety-systems/vaers/index.html. U.S. Department of Health and Human Services. 2024.

[4] Tzu-An Chang et al. “Characterizing and Measuring Linguistic Dataset Drift”. In: arXiv preprint (2023). eprint: 2305 . 17127. url: https://arxiv.org/abs/2305.17127.

[5] Vu Dang et al. “Evaluation of a natural language processing tool for demographic extraction from FDA Adverse Event Reporting System narratives”. In: Frontiers in Drug Safety and Regulation 3 (2022), p. 1020943. doi: 10.3389/fdsfr.2022.1020943.

[6] Jacob Devlin et al. “BERT: Pre-training of Deep Bidirectional Transformers for Language Understanding”. In: arXiv preprint 1810.04805v2 (2019).

[7] OpenAI. Vector Embeddings. 2025. url: https://platform.openai.com/docs/guides/embeddings (visited on 10/23/2025).

[8] Polars: DataFrames for the New Era. https://pola.rs/. Accessed: 2025-09-10.

[9] Nina Tahmasebi, Lars Borin, and Adam Jatowt, eds. Computational Approaches to Semantic Change. Berlin: Language Science Press, 2018. isbn: 978-3-96110-105-3. url: https://langsci-press.org/catalog/book/303.

[10] X. Wang et al. “Active computerized pharmacovigilance using natural language processing and association statistics”. In: Pharmacoepidemiology and Drug Safety 18.6 (2009), pp. 532–540. doi: 10.1002/pds.1744.

